# Towards Treatment Effect Interpretability: A Bayesian Re-analysis of 194,129 Patient Outcomes Across 230 Oncology Trials

**DOI:** 10.1101/2024.07.23.24310891

**Authors:** Alexander D. Sherry, Pavlos Msaouel, Gabrielle S. Kupferman, Timothy A. Lin, Joseph Abi Jaoude, Ramez Kouzy, Molly B. El-Alam, Roshal Patel, Alex Koong, Christine Lin, Adina H. Passy, Avital M. Miller, Esther J. Beck, C. David Fuller, Tomer Meirson, Zachary R. McCaw, Ethan B. Ludmir

## Abstract

Most oncology trials define superiority of an experimental therapy compared to a control therapy according to frequentist significance thresholds, which are widely misinterpreted. Posterior probability distributions computed by Bayesian inference may be more intuitive measures of uncertainty, particularly for measures of clinical benefit such as the minimum clinically important difference (MCID). Here, we manually reconstructed 194,129 individual patient-level outcomes across 230 phase III, superiority-design, oncology trials. Posteriors were calculated by Markov Chain Monte Carlo sampling using standard priors. All trials interpreted as positive had probabilities > 90% for marginal benefits (HR < 1). However, 38% of positive trials had ≤ 90% probabilities of achieving the MCID (HR < 0.8), even under an enthusiastic prior. A subgroup analysis of 82 trials that led to regulatory approval showed 30% had ≤ 90% probability for meeting the MCID under an enthusiastic prior. Conversely, 24% of negative trials had > 90% probability of achieving marginal benefits, even under a skeptical prior, including 12 trials with a primary endpoint of overall survival. Lastly, a phase III oncology-specific prior from a previous work, which uses published summary statistics rather than reconstructed data to compute posteriors, validated the individual patient-level data findings. Taken together, these results suggest that Bayesian models add considerable unique interpretative value to phase III oncology trials and provide a robust solution for overcoming the discrepancies between refuting the null hypothesis and obtaining a MCID.

**SIGNIFICANCE STATEMENT:** The statistical analyses of oncology trials are usually performed by calculating *P* values, although these are poorly understood. Using *P* value cutoffs, such as *P* < 0.05, may lead to some treatments being accepted which have little benefit, and other therapies being rejected which have considerable benefit. A more intuitive and direct probability— that an experimental treatment is better than a standard treatment—can be calculated by Bayesian statistics. Here we used software to obtain the outcomes of 194,129 patients enrolled across 230 trials and then calculated probabilities of benefit. Interpretations based on *P* values disagreed with the probabilities of benefit in one-third of trials. This study suggests that probabilities of benefit would considerably enhance the interpretation of oncology trials.

## INTRODUCTION

The primary endpoints of phase III oncology randomized controlled trials (RCTs) are usually evaluated by frequentist inference.(1) Simplistically, RCTs usually declare superiority of the experimental therapy when *P* is less than the significance threshold for the primary survival endpoint and when directionality is supported by effect estimates, such as the hazard ratio (HR).

Although the published literature is largely based on the frequentist tradition, the use of dichotomized significance thresholds for rule-based decision-making may oversimplify the differences between binary decisions and inferences based on quantitative continuous estimates.(2, 3) In addition to the problems with binary significance thresholds, *P* is frequently misinterpreted by physicians and scientists as the probability that the null hypothesis is true, or alternatively, that 1 – *P* is the probability of superiority for the experimental arm.(4–6) Arguably, this widespread misinterpretation of *P* (and by extension, 95% CI) occurs because physicians and scientists are actually interested in knowing the probability of these hypotheses. Such probabilities quantifying uncertainty are practically more useful to physicians and patients than *P*-values, which represent long-range probabilities of observing more extreme results if the null hypothesis and all auxiliary assumptions were true.(7) Although standard frequentist inference does not provide the probability of hypotheses being true, Bayesian inference can compute these probabilities. Further, within the Bayesian framework, the probabilities for a range of various effect sizes and priors can be determined, allowing important hypotheses beyond the null to be readily examined, such as the probability of clinically meaningful effects.(6)

Thus, we sought to comprehensively re-analyze the primary endpoints of published phase III RCTs in oncology. We reconstructed individual patient-level data from published phase III trials and determined the prevalence of treatment effect misinterpretation by comparing trial interpretation and posterior probabilities (**Fig. S1**). We computed posteriors at various effect sizes, including a minimum clinically important difference (MCID) defined at HR < 0.8. Because current trials are interpreted with respect to HR = 1, we also defined “at least marginal benefit” at HR < 1. To illustrate the potential interpretative value added by Bayesian inference, we compared interpretations of trials based on *P* values versus posterior probabilities. We lastly compared the posteriors of individual patient-level models with those computed by a phase III, oncology-specific prior operative on published summary statistics.(8)

## RESULTS

### Trials and published outcomes

After excluding 24 RCTs that did not meet reconstruction quality criteria and 67 RCTs with proportional hazards violations, 230 two-arm, superiority-design, time-to-event phase III RCTs enrolling 194,129 patients were eligible for analysis (**Fig. S2**). Publication dates ranged from 2005 through 2020. Most RCTs (61%) studied metastatic solid tumors with surrogate primary endpoints (61%) (**Table S1**).

### Summary estimates of the survival function by using Bayesian inference

Bayesian cox regressions were performed on the reconstructed individual patient-level data. The following priors defined on the symmetric scale ln(HR) by the normal distribution *N* (mean, standard deviation) were used: a neutral prior *N* (0, 10^6^), a moderately skeptical prior *N* (0, 0.355), and a moderately enthusiastic prior *N* (–0.44, 0.40) based on published guidelines.(9) As anticipated, the likelihood HRs most strongly favoring the experimental arm seemed to be closer to the null under the skeptical prior compared with the enthusiastic prior (**Fig. S3**). The slope (m) of the ordinary least-squares regression for the mean HR was 0.86 (95% CI, 0.84 to 0.88), which is a quantitative measure of expected shrinkage effects of the skeptical prior. Under the neutral prior, posterior summary estimates were similar to the frequentist results (**Fig. S3**). For example, the median absolute difference between the frequentist HR and posterior mean HR under the neutral prior was 0.001 (max, 0.25).

### Posterior probabilities and trial interpretation

Posterior probabilities under each prior were generally correlated with the published trial interpretation across different hypotheses (**Table S2**).(10) For example, the association between posterior probability of strong clinical benefit under the skeptical prior and trial interpretation of a positive trial was quantified by an aOR of 2.25 (95% CrI, 1.23 to 4.06). The posterior distributions for positive trials were largely identifiable and distinct from the posterior distributions of negative trials (**Fig. 1**).

**Figure 1.**
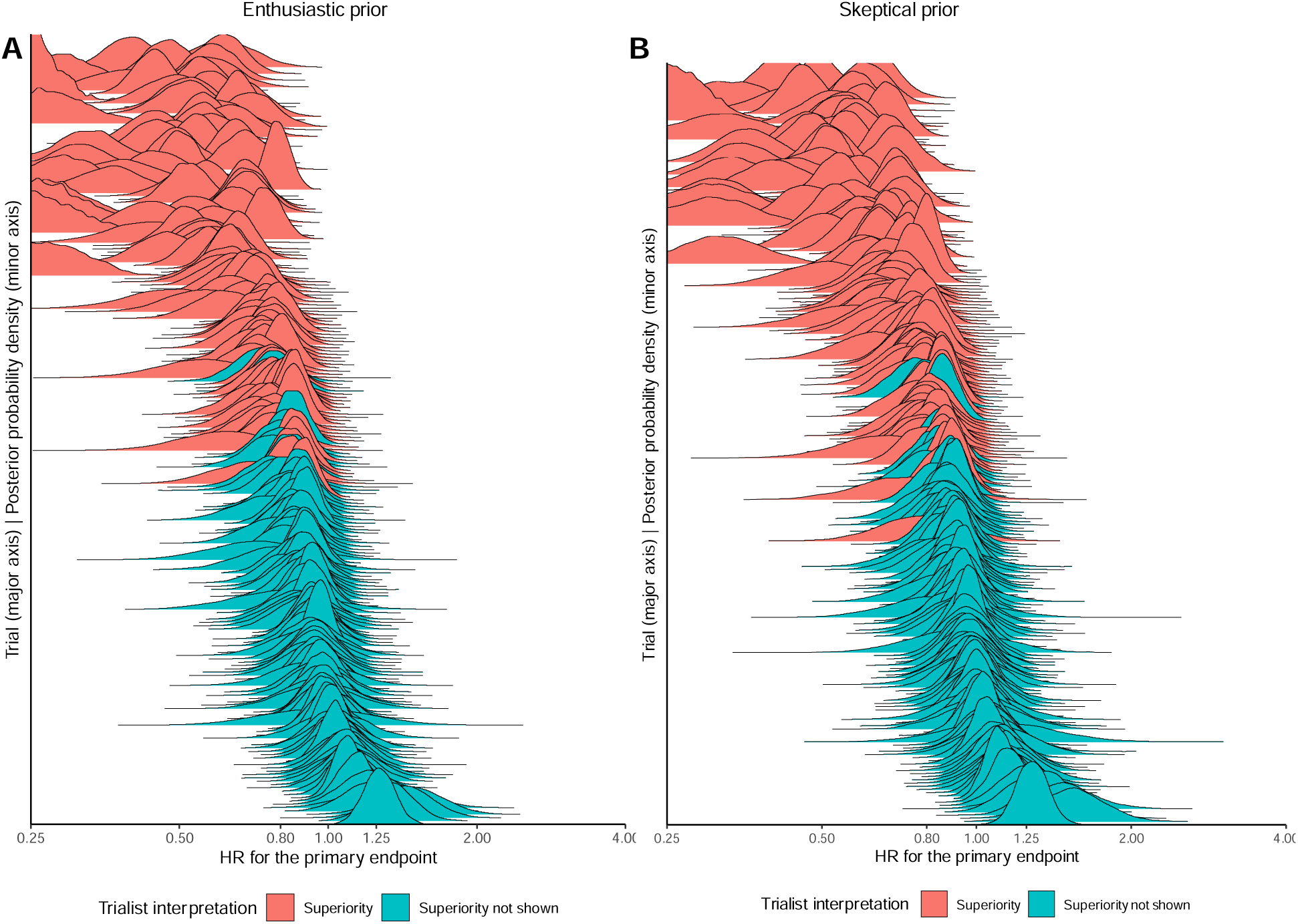
Posterior distributions for all trials plotted in descending order of posterior probability of HR <1 and labeled according to trialist interpretation. Posteriors for the enthusiastic prior are shown in panel A and for the skeptical prior in panel B.

### Posterior probabilities for trials interpreted as positive

All RCTs interpreted as positive had probabilities of achieving at least a marginal benefit exceeding 90% under any prior (**Table 1**). For example, the median posterior probability was 1.00 (IQR, 0.99 to 1.00) under the skeptical prior for HR < 1 (**Table S3**). This finding suggests that posterior probabilities under moderate-strength priors for HR < 1 or HR > 1 replicate the information provided by traditional significance criteria for phase III trials, consistent with the assertions that phase III trials already convey much information in the likelihood distribution and that small values of *P* provide increasingly greater bits of refutational information (increasing *S* values) and confidence in rejecting the null hypothesis (**Fig. 2A and Fig. 2B**).(11)

**Figure 2.**
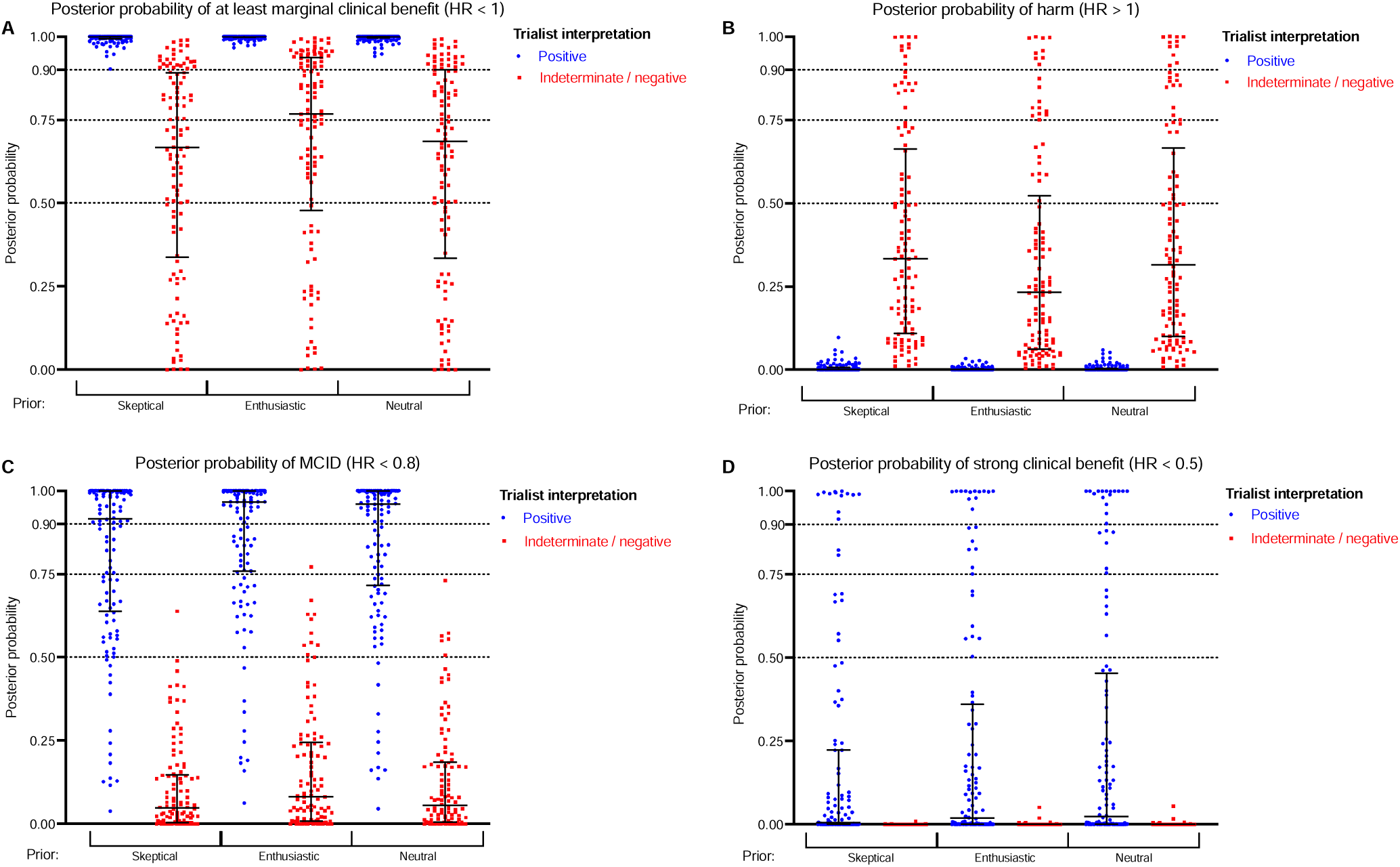
Posterior probabilities under each prior grouped according to trial interpretation. (A) At least marginal clinical benefit hypothesis. (B) Harm hypothesis. (C) MCID hypothesis. (D) Strong clinical benefit hypothesis. Abbreviations: HR, hazard ratio; MCID, minimum clinically important difference.

**Table 1.** Trials with posterior probabilities above 50%, 75%, and 90% are tabulated according to each hypothesis and prior, and grouped by the frequentist interpretation of either positive or indeterminate/negative. Four hypotheses were evaluated: the probability of achieving the minimum clinically important difference (MCID) with the experimental arm, defined as hazard ratio (HR) <0.8; the probability of achieving any benefit with the experimental arm, defined as HR <1; the probability of achieving strong clinical benefit with the experimental arm, defined as HR <0.5; and the probability of harm, HR >1.

| Hypothesis by prior | Posterior probability | According to trial interpretation, n (%) |  |
| --- | --- | --- | --- |
|  |  | Positive (n=120) | Negative (n=110) |
| MCID |  |  |  |
| Skeptical | >90% | 66 (55%) | 0 (0%) |
| Skeptical | >75% | 79 (66%) | 0 (0%) |
| Skeptical | >50% | 106 (88%) | 1 (1%) |
| Neutral | >90% | 73 (61%) | 0 (0%) |
| Neutral | >75% | 86 (72%) | 0 (0%) |
| Neutral | >50% | 109 (91%) | 6 (5%) |
| Enthusiastic | >90% | 74 (62%) | 0 (0%) |
| Enthusiastic | >75% | 92 (77%) | 1 (1%) |
| Enthusiastic | >50% | 110 (92%) | 10 (9%) |
| Strong benefit |  |  |  |
| Skeptical | >90% | 12 (10%) | 0 (0%) |
| Skeptical | >75% | 14 (12%) | 0 (0%) |
| Skeptical | >50% | 20 (17%) | 0 (0%) |
| Neutral | >90% | 16 (13%) | 0 (0%) |
| Neutral | >75% | 22 (18%) | 0 (0%) |
| Neutral | >50% | 27 (23%) | 0 (0%) |
| Enthusiastic | >90% | 13 (11%) | 0 (0%) |
| Enthusiastic | >75% | 20 (17%) | 0 (0%) |
| Enthusiastic | >50% | 27 (23%) | 0 (0%) |
| At least marginal benefit |  |  |  |
| Skeptical | >90% | 120 (100%) | 26 (24%) |
| Skeptical | >75% | 120 (100%) | 48 (44%) |
| Skeptical | >50% | 120 (100%) | 75 (68%) |
| Neutral | >90% | 120 (100%) | 27 (25%) |
| Neutral | >75% | 120 (100%) | 48 (44%) |
| Neutral | >50% | 120 (100%) | 74 (67%) |
| Enthusiastic | >90% | 120 (100%) | 37 (34%) |
| Enthusiastic | >75% | 120 (100%) | 60 (55%) |
| Enthusiastic | >50% | 120 (100%) | 82 (75%) |
| Harm |  |  |  |
| Skeptical | >90% | 0 (0%) | 10 (9%) |
| Skeptical | >75% | 0 (0%) | 20 (18%) |
| Skeptical | >50% | 0 (0%) | 35 (32%) |
| Neutral | >90% | 0 (0%) | 11 (10%) |
| Neutral | >75% | 0 (0%) | 22 (20%) |
| Neutral | >50% | 0 (0%) | 36 (33%) |
| Enthusiastic | >90% | 0 (0%) | 9 (8%) |
| Enthusiastic | >75% | 0 (0%) | 19 (17%) |
| Enthusiastic | >50% | 0 (0%) | 28 (25%) |

Abbreviations: MCID, minimum clinically important difference.

However, the posterior probabilities of achieving the MCID had considerably more overlap between trials interpreted as positive and trials interpreted as indeterminate (**Fig. 2C**). Even under the enthusiastic prior, only 62% of positive RCTs (74 of 120) demonstrated a posterior probability > 90% for achieving the MCID, and 55% of positive RCTs (66 of 120) demonstrated a > 90% posterior probability of meeting the MCID under the skeptical prior (**Table 1**). Thus, even though the experimental arms of RCTs interpreted as positive were likely to have at least marginal benefits compared with the control arm, the probability of a clinically meaningful difference between arms was lower for many RCTs. In turn, because *P* is not a measure of effect size, traditional trial interpretation relying on dichotomized *P* thresholds is at risk of overestimating beneficial treatment effects, as traditional analysis lacks the ability to directly compute the probability of clinically meaningful benefits. The median probabilities of strong clinical benefits, defined as HR < 0.5, were even lower, even under the enthusiastic prior (median, 0.02; IQR, 0 to 0.35) (**Table S3**). Only rare RCTs (11%, 13 of 120) achieved > 90% probability of a strong clinical benefit, even under the enthusiastic prior (**Fig. 2D**). Notably, the primary endpoints of all 13 RCTs with > 90% probability of a strong clinical benefit used surrogate primary endpoints, predominantly progression-free survival (12 of 13, 92%) or time to progression (1 of 13, 8%). Surrogate endpoints may be less clinically relevant than endpoints such as overall survival or quality of life, as has been extensively discussed in the literature.(12–14) No trials demonstrated > 90% probability of a strong clinical benefit for overall survival. Thus, the positive overall survival treatments effects observed in contemporary oncology phase III RCTs over the past two decades have a modest effect size, which, while not surprising, should be interpreted in this light and with respect to their relative toxicity and cost.(3, 15)

We sought to evaluate the incidence of discordance between posterior probabilities computed for the MCID and trial interpretation under the enthusiastic prior. We used the enthusiastic prior for this definition of discordance to minimize the probability of discrepancy with trials, given the meta-analytical nature of our study, and to be as consistent as feasible with the trials’ current interpretation. We defined discordance as a probability ≤ 90% for meeting the MCID under the enthusiastic prior with a positive trial interpretation. With this definition, 46 RCTs (38%) interpreted as positive did not meet the MCID criterion under the enthusiastic prior. The posterior distributions for discordant RCTs are shown in **Fig. S4A** labeled according to the MCID probability. Adjusted logistic regressions using the confounders shown in **Table S4** identified from the directed acyclic graph shown in **Fig. S5** suggested that the only trial-level factor correlated with discordance between posterior probability and trial interpretation was the use of an overall survival primary endpoint (versus surrogate endpoint, aOR, 2.05; 95% CrI, 1.21 to 3.49) (**Table S5**). Of the 46 discordant RCTs, the primary endpoint was overall survival for 21 RCTs (46%) and surrogate endpoints for 25 RCTs (54%). Because of several differences between surrogate and overall survival endpoints, such as the likelihood of a positive finding, clinical and regulatory interpretation, and time to publication, we performed a subgroup analysis for RCTs with a primary endpoint of overall survival.(16–19) The posterior distributions for RCTs with overall survival as the primary endpoint are shown in **Fig. S4B**. The percentage of RCTs interpreted as positive for overall survival with a posterior probability for the MCID > 90%, > 75%, and > 50% under the enthusiastic prior was 30% (9 of 30), 47% (14 of 30), and 77% (23 of 30), respectively (**Table S6**). The median posterior probability of meeting the MCID for overall survival among RCTs interpreted as positive under the enthusiastic prior was 0.73 (IQR, 0.58 to 0.96) (**Table S7**).

Lastly, we repeated the analysis in the subset of positive trials which led to Federal Drug Administration regulatory approval.(16) Of 120 positive trials overall, the data from 82 trials led to regulatory approval. The median posterior probability of meeting the MCID under the enthusiastic prior was 0.99 (IQR, 0.84 to 1.00). Among these trials, 30% (25 of 82) had ≤ 90% probability for meeting the MCID criterion under the enthusiastic prior, suggesting potential discordance with the trial and regulatory interpretation.

### Posterior probabilities for trials interpreted as negative

The posterior probabilities of at least marginal benefit and harm for RCTs interpreted as negative varied widely under each prior (**Fig. 2A-B**). The median probability of marginal benefit was 0.67 (IQR, 0.32 to 0.89) under the skeptical prior and 0.77 (IQR, 0.49 to 0.94) under the enthusiastic prior (**Table S3**). For these RCTs interpreted as having no advantage with the experimental arm, Bayesian models under each prior, contrary to trial interpretation, suggested that the primary outcome for the experimental arm was more likely superior to the control arm than not (i.e., > 50% posterior probability) in at least 63% of negative RCTs, and up to 75% of negative RCTs based on the prior (**Table 1**).

We defined discordance between RCTs interpreted as negative and posterior probability as posterior probability > 90% for at least a marginal benefit under the skeptical prior.

We used the skeptical prior for this definition of discordance to be as consistent as feasible with the trials’ current interpretation, and we used the hypothesis HR < 1.0 because frequentist interpretations of no difference are specifically regarding the null hypothesis of HR = 1.0. The posterior probability of at least marginal benefit exceeded 90% in 24% of RCTs interpreted as negative (26 of 110) (**Fig. S6A**). More recently published RCTs seemed less likely to have underestimation of treatment effects (aOR, 0.83; 95% CrI, 0.69 to 0.98) (**Table S8**). Of these 26 discordant RCTs, 12 RCTs had a primary endpoint of overall survival (**Fig. S6B**). On February 12, 2024, we queried the National Comprehensive Cancer Network guidelines to evaluate whether the experimental arms of the 12 discordant RCTs, interpreted as negative but with > 90% probability of at least a marginal overall survival benefit (HR < 1), were listed as potentially useful regimens in the clinical setting in which they were tested. Of the 12 trials, 1 regimen was the preferred strategy; 3 regimens were considered options or useful in certain circumstances, and the remaining 8 regimens were not listed in the guidelines (67%).

Consistent with the notion that large values of *P* (i.e., *P* > 0.05) provide a low degree of refutational evidence, the median *S* for discordant RCTs was 3 (IQR, 2 to 4), a result as surprising as a coin toss achieving tails on three consecutive tosses (**Fig. S7**). As a reference, the median *S* for positive RCTs was 12 (IQR, 7 to 21), and the median *S* for positive RCTs without discordance was 19 (IQR, 12 to 28). In total, these findings suggest that frequentist analyses, when used in isolation, often result in underestimation of treatment effects and potential rejection of superior therapeutic strategies.

### Comparison of posterior probabilities to a phase III, oncology-specific prior

Because of the limitations of reconstructed data, we sought to validate these findings by also computing posteriors based on the summary statistics for each primary endpoint, using a phase III, oncology-specific prior leveraging an exchangeable conjugate approach.(8) This phase III, oncology-specific prior was derived based on the treatment effect distribution of 415 phase III trials, inclusive of the 230 trials analyzed in the present study, as previously reported.(8)

Posterior estimates, derived from summary statistics using the phase III, oncology-specific prior, were concordant with posterior estimates computed by the conventional priors using the reconstructed individual patient-level data (**Fig. 3**). For example, the concordance coefficient between the posterior probability of the MCID under the skeptical prior versus the phase III, oncology-specific prior was 0.98 (95% CI, 0.98 to 0.99) (**Table S9**). This finding provides additional validation for posteriors computed by the phase III, oncology-specific prior.

**Figure 3.**
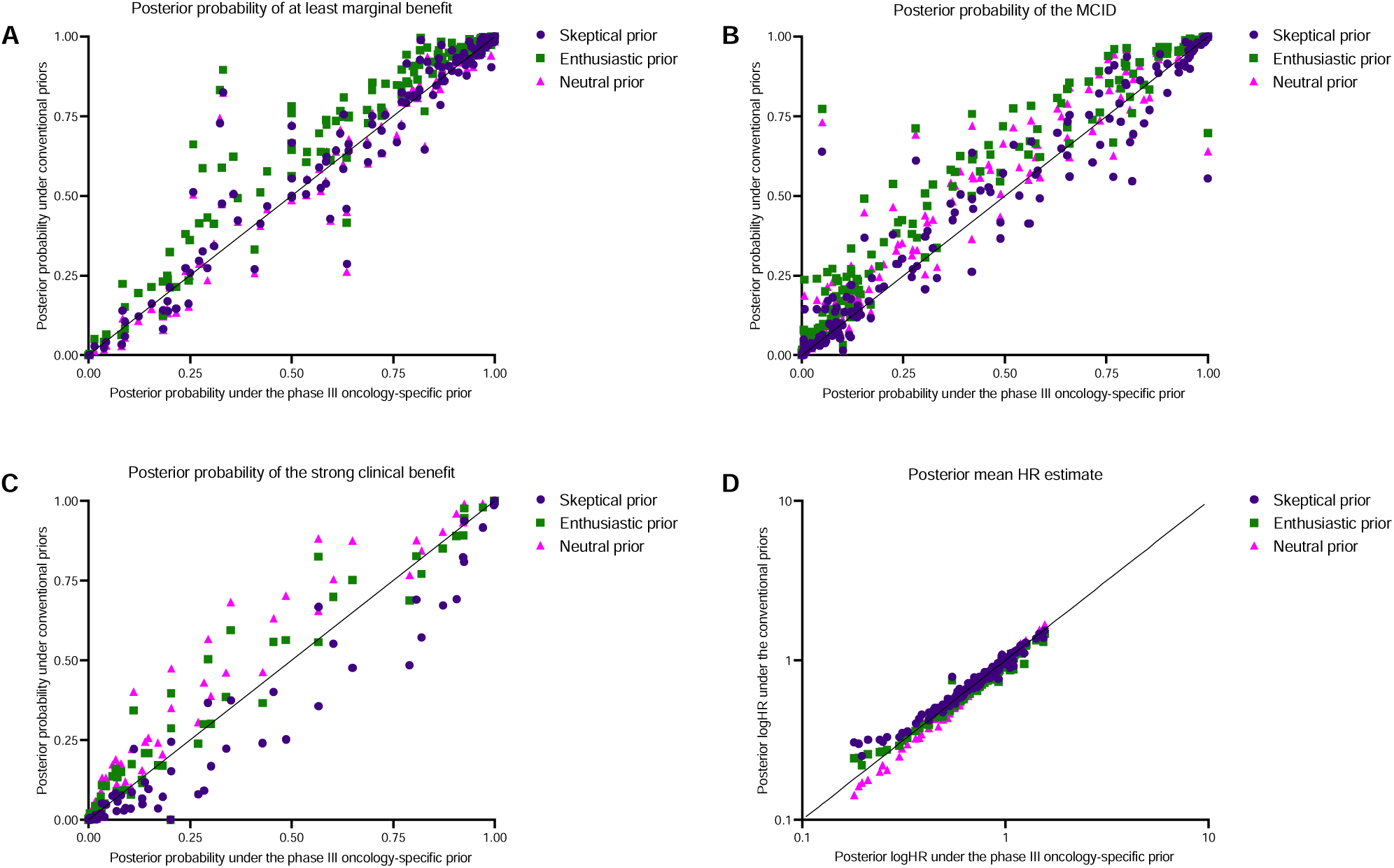
Comparison of posterior outputs from individual patient-level data using conventional priors versus summary statistics using a phase III, oncology-specific prior. A line of identity has been added. (A) Posterior probability of at least marginal benefit (HR < 1). (B) Posterior probability of achieving the MCID (HR < 0.8). (C) Posterior probability of strong clinical benefit (HR < 0.5). (D) Posterior mean estimate of HR.

## DISCUSSION

In this large-scale analysis of 230 trials, Bayesian inference added unique, complementary, and intuitive interpretative value towards understanding the primary results of phase III oncology RCTs. Trials interpreted as positive had low probabilities of MCID in 38% of cases, and trials interpreted as negative had high probabilities of at least marginal differences in 24% of cases. The refutational support was low for RCTs interpreted as negative but associated with high probabilities of benefit. Taken together, these findings suggest that current approaches and overreliance on binary *P* thresholds for significance may result in overestimation and underestimation of relevant treatment effects. Posterior probabilities should be included in the analyses of future phase III oncology RCTs to facilitate more intuitive, clinically meaningful, and robust trial interpretations.

This study provided an empirical assessment of the extent to which the information loss induced by dichotomization into binary significance thresholds may lead to the under- or over-estimation of effects in phase III oncology RCTs. Overestimation of benefit was demonstrated in a substantial proportion of trials, even with an enthusiastic prior. Changing the focus of trial interpretation from information against the null hypothesis (the typical use of *P*) towards the probability of achieving a clinically meaningful effect size may better harmonize the gap between clinical trial results and real-world efficacy, spare patients costly and toxic therapies associated with only marginal benefits, and guide interpretation of results to the broader community including patients, regulatory agencies, lay press, and more.(6, 15, 16, 20, 21) We also observed underestimation of benefit, even with use of a skeptical prior.(22) This limitation of frequentist inference was exemplified by the discordance for trials interpreted as negative, all of which had low refutational information (lower *S* values / larger *P* values). Finally, although re-analyses of several select RCTs have illustrated the interpretative value added by Bayesian inference, none are of the scope and scale of the present study.(23–25) Collectively, this study provides strong and pragmatic findings of the potential consequences of misinterpreting *P*-values greater than 0.05 as evidence of no difference and small *P*-values as evidence of clinically meaningful difference.(26–28)

A barrier to the uptake of Bayesian inference in oncology has been the requirement to specify the prior probability of the hypothesis.(29, 30) While some have argued that the main disadvantage of Bayesian inference is potential bias of the posterior from prior selection, incorporation of prior probability is one of the chief advantages of Bayesian inference. Pre-specification of the prior makes explicit the research assumptions and knowledge preceding the study, and allows formal evaluation of differences in opinion.(31) For phase III trials, as our data show, moderate-strength priors also largely preserved the likelihood distribution in the posterior. This feature may be particularly appealing in settings in which there is concern for subjectivity in the choice of the prior or in which there is concern that the prior may devalue interpretation of the posterior by producing starkly different results from frequentist inference. Furthermore, numerous subjective decisions are also made in frequentist RCTs regarding experimental design, power estimation and pre-specification of expected treatment effect size, α thresholds, and stopping rules.(32, 33) These decisions can heavily influence results, and should not be regarded as different in kind from specifying a prior. Ideally, prior(s) are developed by content-matter experts in conjunction with statisticians familiar with Bayesian methods at the time of trial design. By leveraging prior information, RCTs with sequential Bayesian designs may be more efficient and require fewer patients for testing the same hypotheses as frequentist RCTs.(34, 35) The phase III, oncology-specific prior may be suitable for physicians or others who are interested in estimates of specific published trials, and they can be computed by using a simple online calculator provided by our previous study without the need for reconstruction or using software code.(8) In the absence of strong domain-specific prior knowledge, based on the validation provided by the present study, the phase III, oncology-specific prior is a reasonable default informative prior for phase III oncology trials.

This study had several key limitations. First, potential discordance was defined by discrepancies between the frequentist interpretation and the Bayesian conclusion at various thresholds of posterior probability. Although a landmark Bayesian re-analysis of critical care RCTs used posterior probability thresholds of 50% to define potential benefit, a more conservative threshold was selected here based on intrinsic differences between the fields of critical care and oncology and the greater heterogeneity in primary endpoints in oncology compared with critical care.(24) However, the full posterior probability distribution would be more informative to communicate in an individual trial than dichotomized probability thresholds. The approach used here should not imply that thresholds exist for translating Bayesian inference into making clinical decisions, which rely on numerous factors specific to the clinical scenario.(3, 15) The re-analysis findings of potential efficacy in negative RCTs should not be taken as suggestions to alter current clinical decision-making, especially in the absence of individual patient-level data for each study, explicit conditioning on prognostic variables in the regression models, and assessment of other modeling assumptions. Second, the MCID was set uniformly for all studies encompassing several different disease settings, which may be an oversimplification. The MCID may depend on the specific disease setting and prognosis, and definitions for the MCID often include both absolute and relative differences in outcomes.(20) Third, this study focused on the primary endpoint, not taking into account other relevant secondary endpoints, such as patient-reported outcomes, toxicity, or secondary efficacy analyses, that may affect application of trial results to clinical practice. Ideally, efficacy and safety would be evaluated simultaneously, at the individual patient level, by defining a primary composite endpoint that takes both into account.(36) Primary endpoints differed between trials (overall survival vs surrogate survival) which may have further affected the findings, and likely the effect of surrogate survival endpoints are larger than overall survival effects. Trial setting, including whether the control arm was placebo vs an active comparator, may have further influenced the data. Fourth, use of reconstructed data, which served as the basis for the Markov Chain Monte Carlo models, has several inherent limitations.(37) In addition, the reconstructed data were univariable and unadjusted in nature, as they were derived from Kaplan-Meier curves. A lack of adjustment for prognostic covariates may have limited the efficiency and increased the uncertainty of the Bayesian models compared with the Cox regressions computed by the trials, although the phase III, oncology-specific conjugate prior was not subject to the reconstruction limitation and not necessarily subject to the univariable limitation, as it was instead based directly on the underlying trial regression model. Proportional hazards violations in the reconstructed data were observed in a fair number of trials; because this may have influenced estimates for all models, these trials were excluded from further analysis, although this may have affected overall findings.

In summary, the present empirical Bayesian assessment of the primary endpoints of published phase III oncology RCTs suggests that treatment effects are often either underestimated or overestimated with traditional analyses. Computing Bayesian posterior probabilities, either using individual patient-level data or the summary statistics, is an appealing strategy to mitigate the misinterpretation of trial results based on *P* values and to provide more intuitive and robust evaluations the chances that a treatment is clinically meaningful.

## ONLINE METHODS

### Design and Inclusion

Institutional review board approval for this cross-sectional study was not needed because of the public availability of trial data. Trials were identified from ClinicalTrials.gov based upon previously defined search criteria.(38) Phase III, two-arm, superiority-design, oncology trials testing therapeutic anti-cancer strategies with a time-to-event primary endpoint were included. Ongoing trials were excluded if the final analysis of the primary endpoint analysis had not yet been published. If 95% CI was used for all time-to-event co-primary endpoints, overall survival was evaluated based on its interpretability and potential advantages compared with surrogate endpoints.(1, 2) This study conforms with the recommendations of the Strengthening the Reporting of Observational Studies in Epidemiology (STROBE) guidelines and the reporting of Bayes used in clinical studies (ROBUST) guidelines for Bayesian analysis.(39, 40)

For each trial, the results for the primary endpoint for were recorded, including the published interpretation of the trial primary endpoint (i.e., significantly better with the experimental therapy or not according to the trial’s pre-specified criteria), the HR, the 95% CI, and the *P*-value. To facilitate analysis, in trials where the experimental arm was taken as the reference group, reciprocals of the HR and 95% CI were taken to recast the control arm as the reference group. All survival time units were harmonized to months.

### Reconstruction of individual patient-level data

After trials were screened from ClinicalTrials.gov, individual patient-level data were reconstructed from the Kaplan-Meier curves of the primary endpoint for each trial by using WebPlotDigitizer and established reconstruction methods.(37) The quality of the reconstruction was determined by performing a Cox regression on the reconstructed data and then calculating the absolute value of the natural logarithm of the ratio for the HR of the reconstructed data and the HR reported in the manuscript. High-quality reconstructions were defined by values ≤ 0.1, and reconstructions meeting this metric were eligible for further analysis. Proportional hazards were assessed in each high-quality reconstruction by using previously described methods.(41)

### *S* computation when *P* was not provided

The information against the null hypothesis based on frequentist inference was defined as the surprisal (*S*) value. Calculated as –log_2_(*P*), *S* encodes the number of bits of refutational information, and may be more intuitive than *P* values.(2, 11) *S* can be conceptualized as how surprising the observed data would be if the null hypothesis were true. Rounded to the nearest integer, *S* represents the number of consecutive coin flips of a fair coin toss that would need to land on the same side to confer an equivalent level of surprise. For example, *S* = 5 indicates that, under the null hypothesis, the probability of observing a result as or more extreme is as surprising as always observing tails in 5 consecutive tosses of a fair coin. We computed *S* values for each trial from reported *P* values by using the calculator provided by Msaouel and colleagues.(2) For RCTs where a range of *P*-values was provided in lieu of a specific *P* (e.g., *P* < 0.001), *P* values were back-computed by established approaches from Altman and Bland by using the calculator provided by Msaouel and colleagues.(2, 42) One-sided *P* values were converted to two-sided *P* values based on the direction of effect observed by either multiplying the reported *P* by two if benefit was observed (HR < 1) or subtracting the *P* from 1 and then multiplying by two, because all RCTs hypothesized superiority of the experimental arm versus the control arm.

### Bayesian survival analysis

Bayesian cox regressions were performed on the reconstructed individual patient-level data. The following priors defined on the symmetric scale ln(HR) by the normal distribution *N* (mean, standard deviation) were used: a neutral prior *N* (0, 10^6^), a moderately skeptical prior *N* (0, 0.355), and a moderately enthusiastic prior *N* (–0.44, 0.40) based on published guidelines.(9) Moderate-strength priors were chosen based on the assumption that a reasonable body of evidence existed in support of the experimental therapy before the initiation of most phase III RCTs. Markov chain Monte Carlo sampling was used with 4 chains, 2000 burn-in steps, and 8000 total iterations by using R v4.3.2 with the package brms.(43, 44) Convergence was assessed by calculating effective sample size and R-hat, with convergence defined by an effective sample size ≥1000 and R-hat <1.05.(44) All Bayesian models included in this study met convergence criteria.

Posterior probabilities were also computed based on the published Cox regression summary statistics for each trial (HR and 95% CI) using a phase III, oncology-specific prior as described previously.(8) The treatment effect distribution was obtained based on previously reported methods used to derive a shrinkage estimator from RCTs in the Cochrane database; in brief, the z-score distribution was deconvoluted into the signal-to-noise ratio distribution, which was then scaled by the standard error to obtain the treatment effect prior probability distribution.(45–49)

### Clinically relevant hypotheses

Although frequentist and Bayesian inferences are complementary approaches, a principal advantage of Bayesian inference is the direct determination of probabilities for parameter values such as HRs. We computed posterior probabilities for four clinically relevant questions outlined here: based on criteria from the American Society of Clinical Oncology Cancer Research Committee and others, the minimum clinically important difference (MCID) was defined as 1) HR < 0.8.(3, 20, 50) Strong clinical benefit was defined as 2) HR < 0.5, at least marginal benefit was defined as 3) HR < 1, and harm was defined as 4) HR > 1.(51)

### Statistical analysis

Continuous variables were summarized by using the median and IQR. Categorical variables were summarized by using the incidence and frequency. Correlations between frequentist and posterior summary estimates were assessed by Lin’s concordance correlation coefficient (ρ_c_). The associations between trial-level covariates and discordance between trial interpretation and posterior probabilities were assessed by Bayesian adjusted logistic regression models, yielding aOR and 95% CrI. Confounders for adjusted Bayesian regressions were identified by structural causal models depicted on directed acyclic graphs by using DAGitty (**Fig. S5**).(52) Each variable was sequentially rotated as the predictor of interest in the directed acyclic graph to select predictor-specific confounders, consistent with prior work.(38) The list of confounders for each predictor are shown in **Table S4**. Adjusted Bayesian logistic regressions were also used to evaluate the association of posterior probability with trial interpretation, for which industry sponsorship and primary endpoint type (surrogate vs overall survival) were selected as confounders based on the directed acyclic graph informed by prior studies (**Fig. S8**).(16, 17, 19, 53) The skeptical prior was used for the logistic models, and the same Markov chain Monte Carlo parameters from the Cox regressions were applied for the logistic regressions. Plots were made in R or Prism v10 (GraphPad, La Jolla, CA).

## Data availability

Reconstructed data used for this study have been uploaded to the data repository figshare and may be accessed at the following webpage: https://figshare.com/articles/dataset/Reconstructed_survival_data_from_Phase_3_oncology_trials/26103268?file=47244859.

## Acknowledgments

We thank Christine Wogan, MS, ELS, of The University of Texas MD Anderson Cancer Center for editing the manuscript. She received no compensation beyond her salary for this contribution. We thank Dr. Erik van Zwet of Leiden University Medical Center for helpful discussions on the content of the manuscript.

## Contributions

A.D.S., P.M., G.S.K., T.A.L., Z.R.M., and E.B.L. conceptualized and designed the study. A.D.S., G.S.K., T.A.L., J.A.J., R.K., M.E.A., R.P., A.K., C.L., and E.B.L. curated the data. A.D.S. and Z.R.M. did the formal analyses. E.B.L. acquired funding. P.M. and E.B.L. supervised the study and provided study resources. A.D.S. prepared the visualizations. All authors contributed to the interpretation of the data. A.D.S. wrote the first draft, and all authors reviewed and revised the manuscript. All authors had full access to all the data in the study and accept responsibility to submit for publication.

