## Supplement for "Towards Treatment Effect Interpretability: A Bayesian Re-analysis of 194,129 Patient Outcomes Across 230 Oncology Trials"

[Figure S7. Comparison of S values versus posterior probability of at least marginal benefit (HR <1) for trials interpreted as negative.	9](#_Toc168829512)

### **Figure S1**. Study schema of patient-level data reconstruction and Bayesian cox regressions under three priors (skeptical) to compute posterior distributions and probabilities for clinically relevant hypotheses.

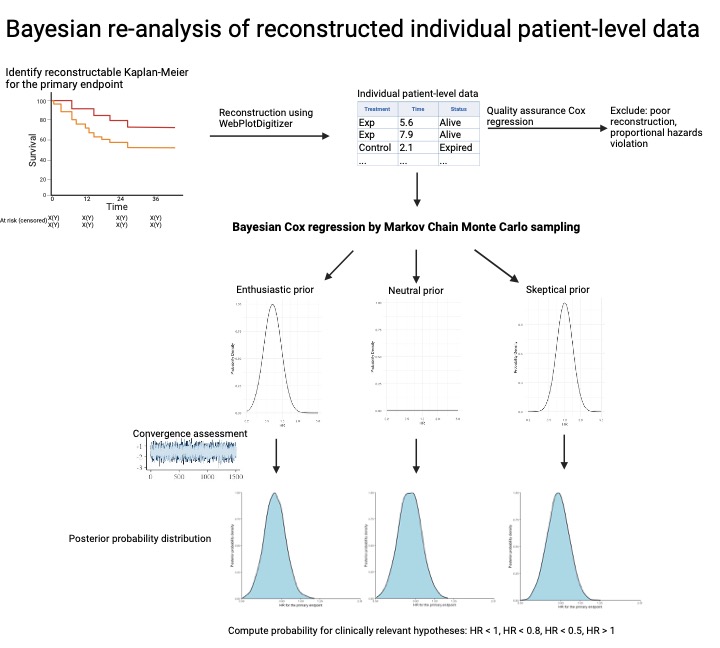

### **Figure S2**. Trial selection flowchart.

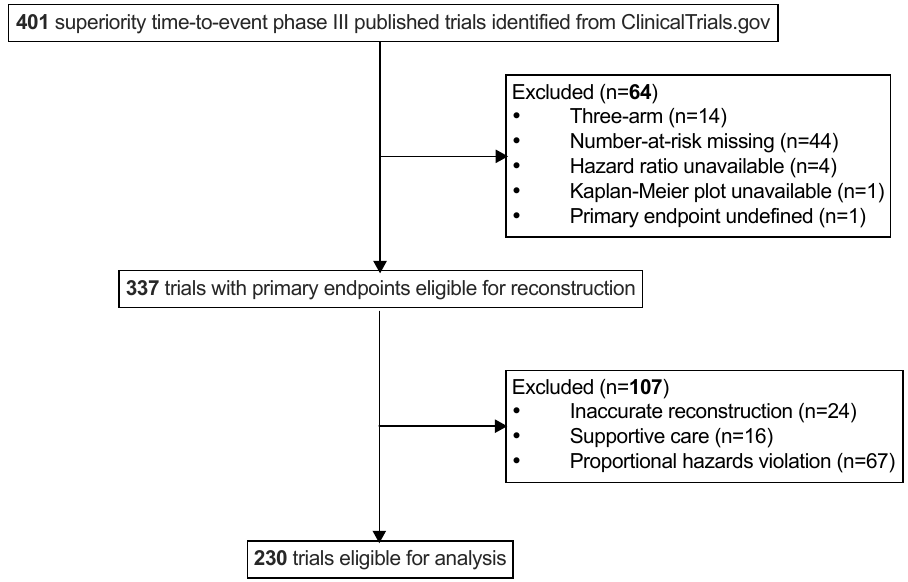

**Figure S3**. Comparison between the frequentist likelihood hazard ratio (HR) estimates and Bayesian posterior HR estimates. A line of identity has been drawn from the origin through the center of the plot for visualization of the prior effects above and below the null (HR = 1). The x-axis and y-axis are transformed to the logarithmic scale for the HR and 95% interval comparisons, and are linear for the interval width comparisons.

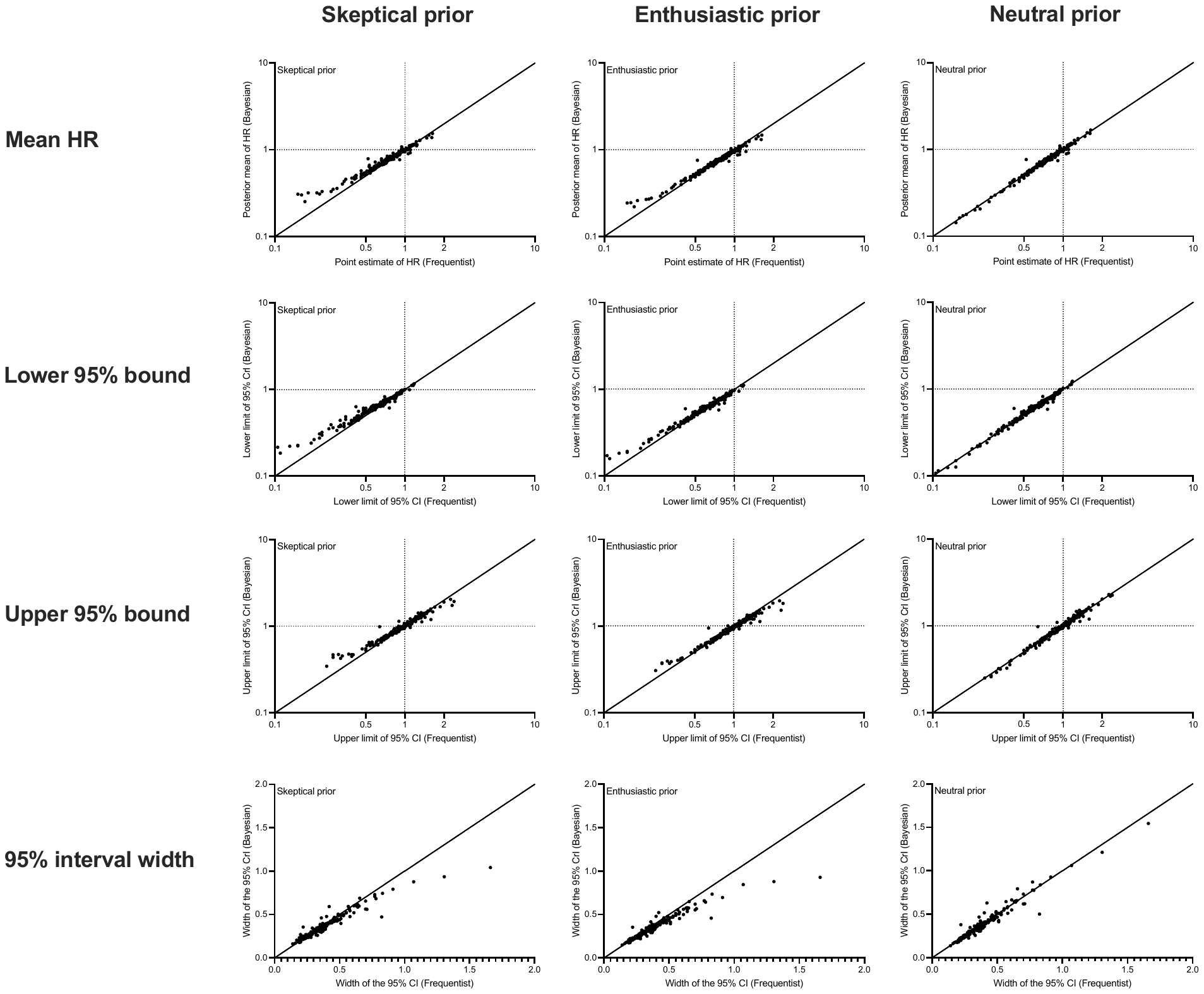

**Figure S4.** Posterior probability distributions of positive trials with potential discordance under the enthusiastic prior. Each trial’s distribution is labeled according to the posterior probability of achieving the MCID. Panel A shows all positive trials with probability ≤90% of achieving the MCID, and panel B shows all positive trials for overall survival with probability ≤90% of achieving the MCID.

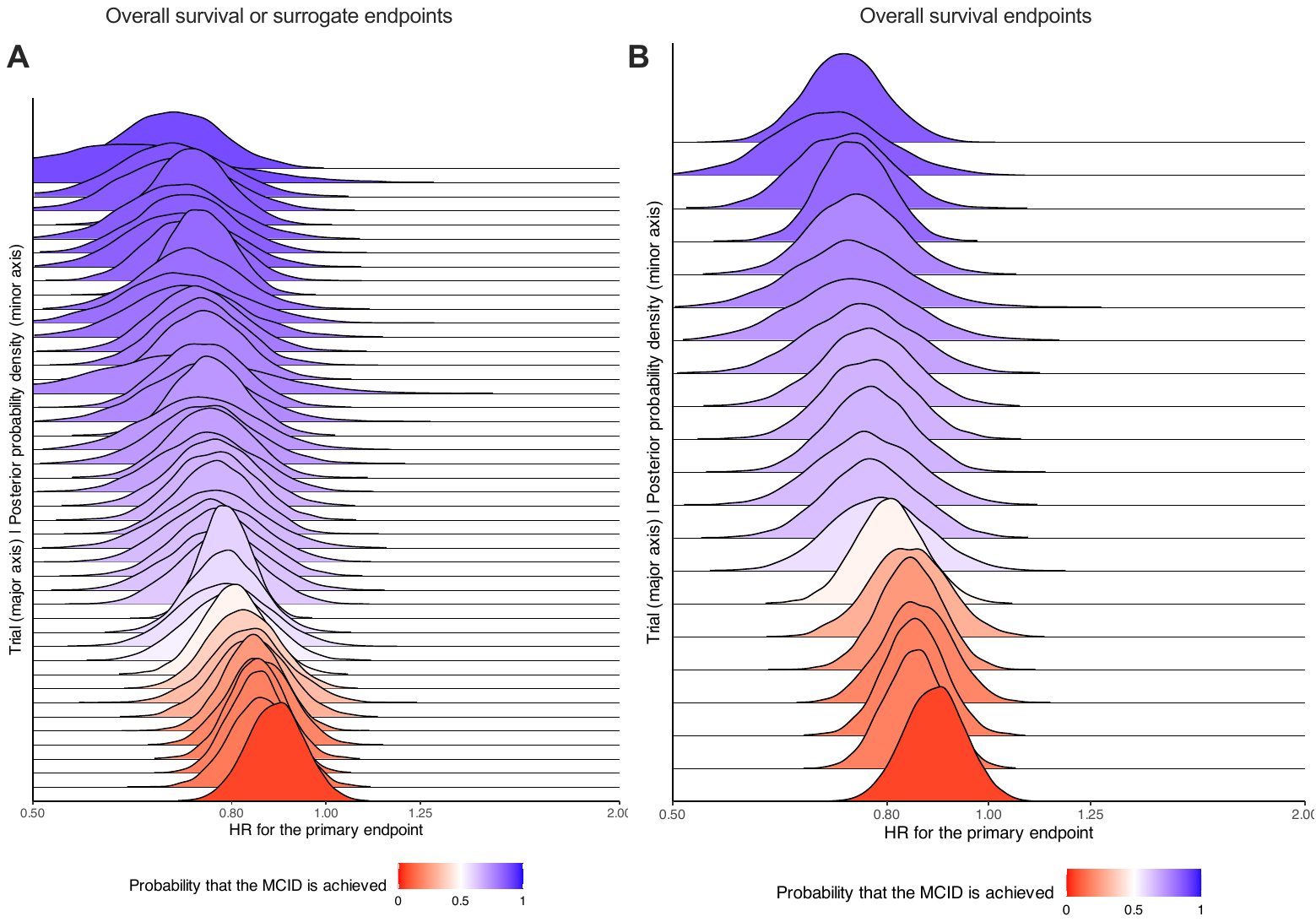

**Figure S5.** Directed acyclic graph for the structural causal model used to identify confounders for the outcome of discordance. In this example, discordance (shown in blue) is the outcome and primary endpoint category (shown in green) is the predictor. The green arrows represent the causal pathways. Confounders for the association between the predictor and outcome are shown in yellow circles.

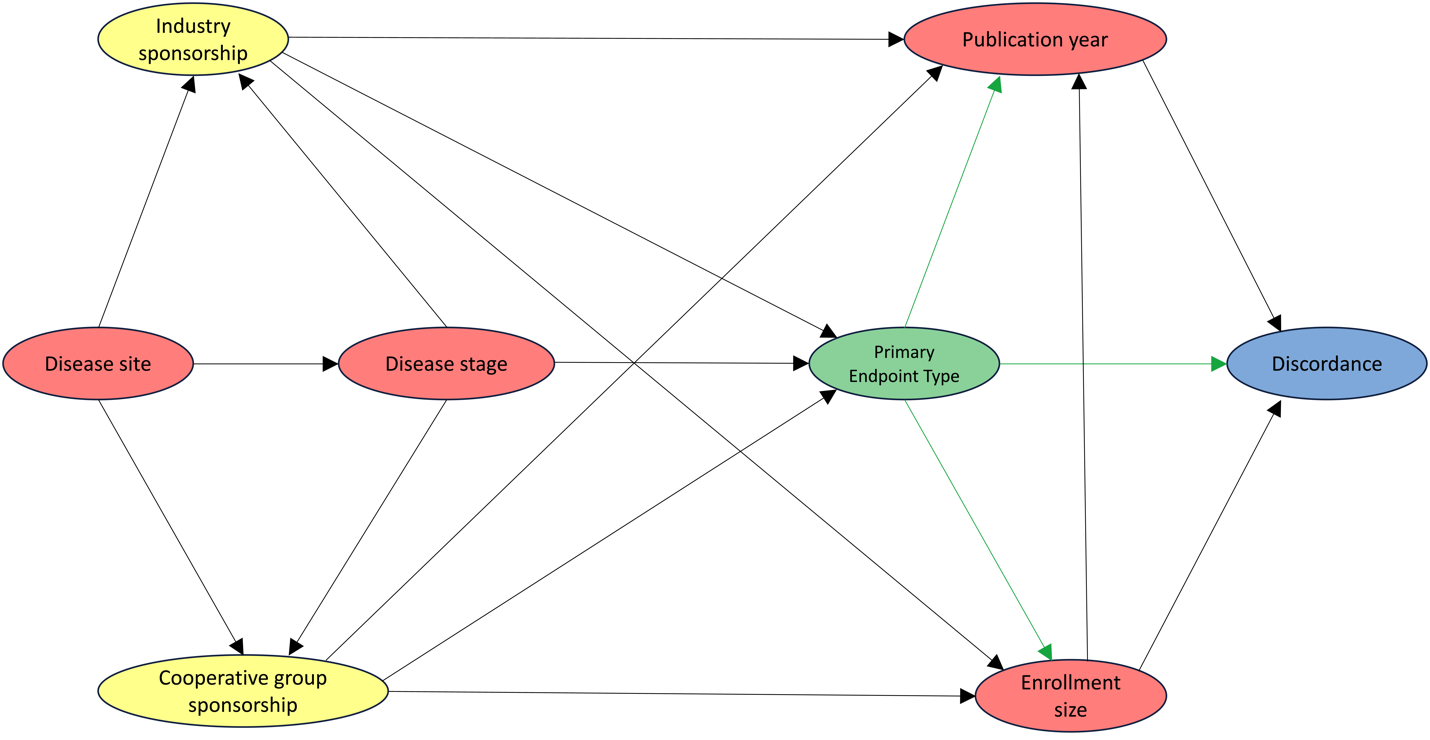

**Figure S6**. Posterior probability distributions of negative trials with potential discordance under the skeptical prior. Trials included are those interpreted as negative by frequentist inference and with posterior probability >90% of at least a marginal benefit. Each trial’s distribution is labeled according to the posterior probability of achieving HR <1. Panel A shows all negative trials with potential discordance, and panel B shows all negative trials for overall survival with potential discordance.

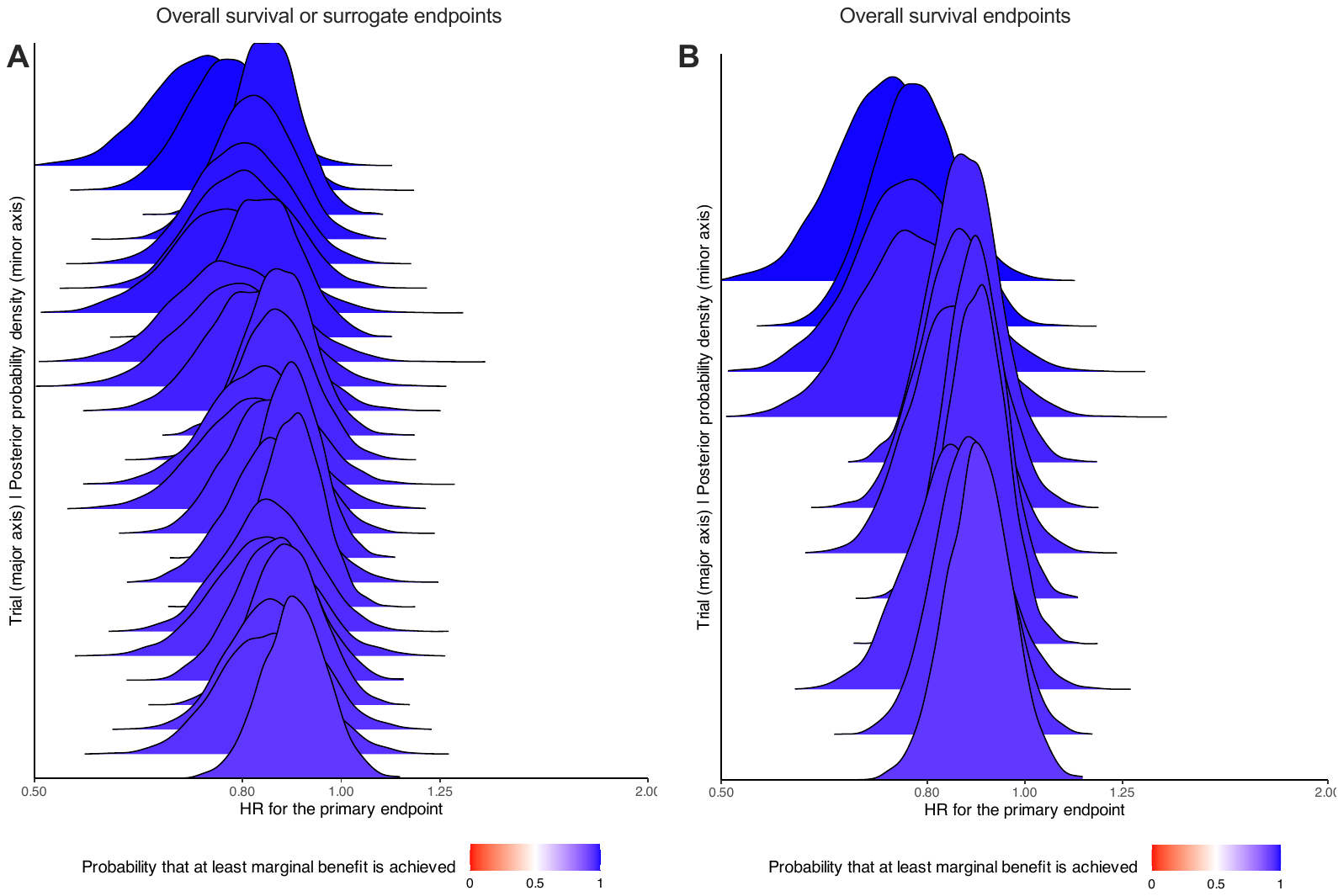

**Figure S7.** Comparison of S values versus posterior probability of at least marginal benefit (HR <1) for trials interpreted as negative. Trials with potential discordance at >90% posterior probability of HR <1 under the skeptical prior are highlighted in red. Trials with high *S* with low probability of benefit were claimed to show inferiority of the experimental therapy.

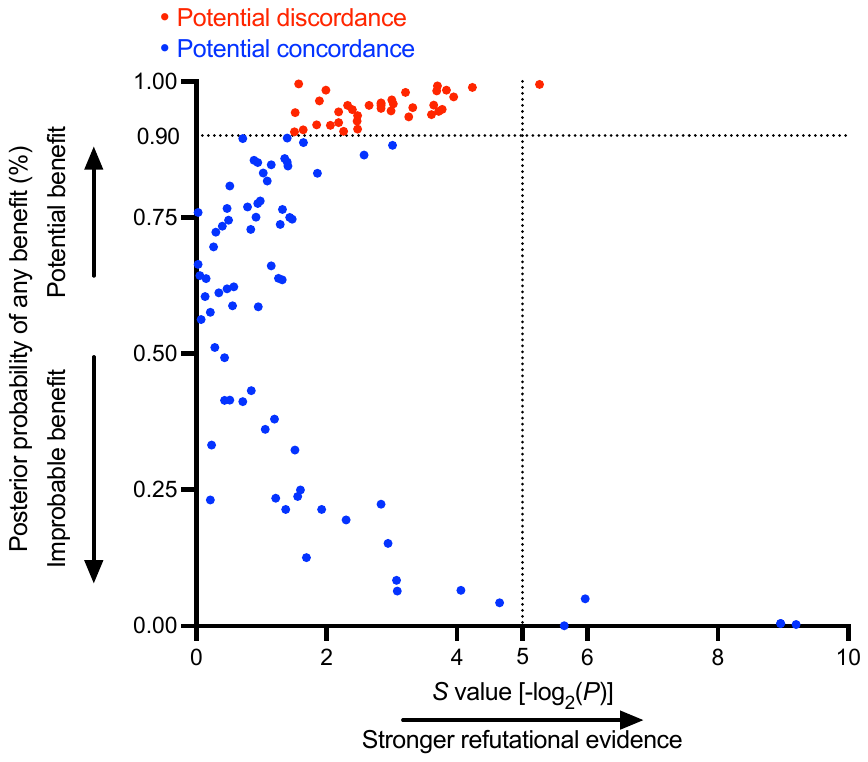

**Figure S8.** Directed acyclic graph for the structural causal model used to identify confounders for the outcome of trialist interpretation. The green arrows represent the causal pathways. Confounders for the association between the predictor (posterior probability) and outcome are shown in yellow circles.

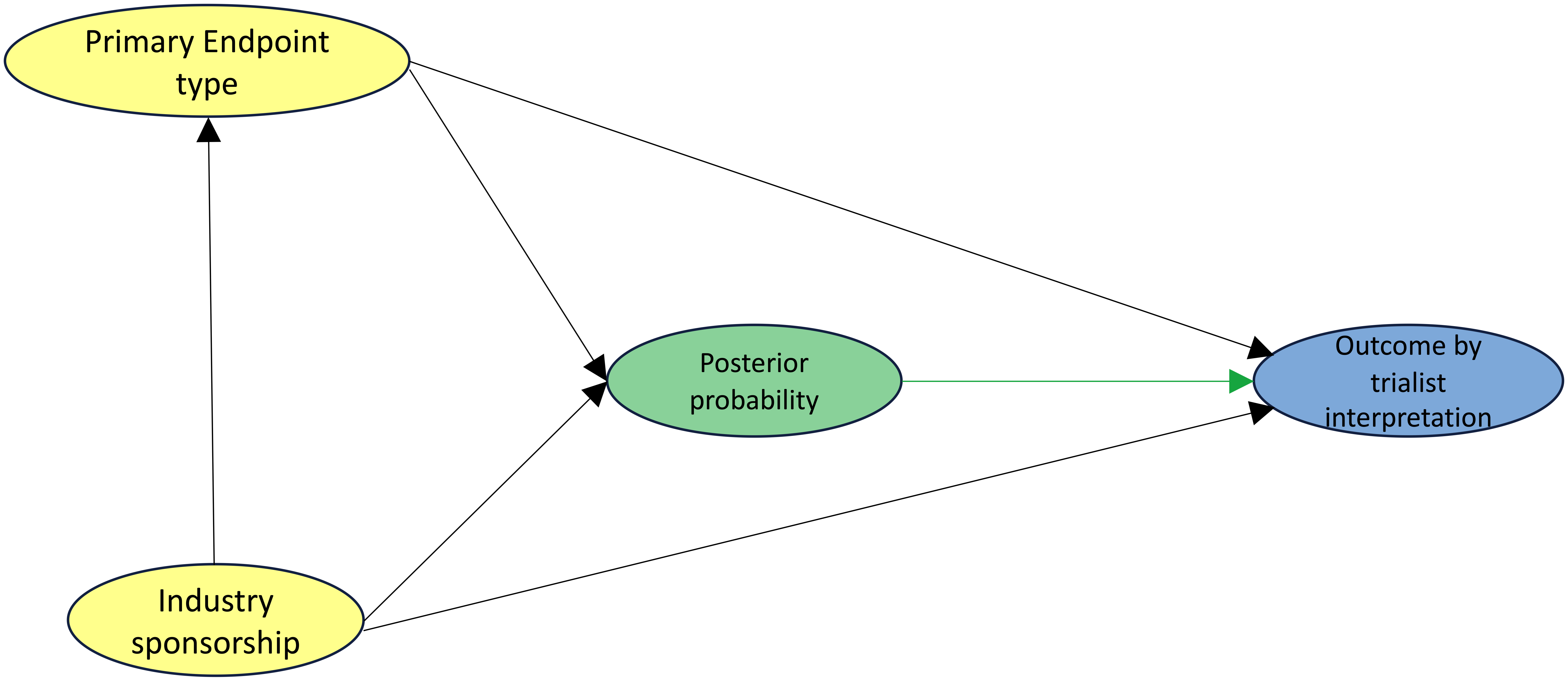

### **Table S1**. Characteristics of trials included in the analysis.

| **Characteristic** | **Value** |
| --- | --- |
| Disease stage, n (%) |  |
| Solid M0 | 51 (22) |
| Solid M1 | 141 (61) |
| Hematologic | 38 (17) |
| Disease site |  |
| Breast | 45 (20) |
| Gastrointestinal | 41 (18) |
| Genitourinary | 35 (15) |
| Hematologic | 38 (17) |
| Thoracic | 40 (17) |
| Other* | 31 (13) |
| Treatment modality, n (%) |  |
| Systemic therapy | 224 (97) |
| Local therapy | 6 (3) |
| Cooperative group sponsorship, n (%) | 53 (23) |
| Industry sponsorship, n (%) | 202 (88) |
| Number of enrolled patients, median (IQR) | 640 (424 to 904) |
| Publication year, median (IQR) | 2015 (2013 to 2017) |
| Primary endpoint, n (%) |  |
| Overall survival | 90 (39) |
| Surrogate** | 140 (61) |
| Primary outcome, n (%) |  |
| Improved efficacy | 120 (52) |
| No improvement in efficacy | 105 (46) |
| Worse efficacy | 5 (2) |

Abbreviations: solid M0, nonmetastatic solid tumor; solid M1, metastatic solid tumor; IQR, interquartile range.

*Other disease sites include central nervous system, endocrine, gynecologic, head and neck, and skin.

**Examples of surrogate endpoints include progression-free survival, disease-free survival, and event-free survival.

**Table S2.** The association of posterior probability and the interpretation of a positive trial. Multivariable Bayesian logistic regressions were used with a skeptical prior adjusted for the confounding covariates of industry funding and primary endpoint category (overall survival vs surrogate endpoint). The MCID hypothesis was defined as HR <0.8, strong clinical benefit was defined as HR <0.5, and at least marginal benefit was defined as HR <1.

| **Posterior by hypothesis and prior** | **Positive Trialist Interpretation** | |
| --- | --- | --- |
|  | **aOR** | **95% CrI** |
| MCID |  |  |
| Skeptical | 10.59 | 6.30 to 17.99 |
| Enthusiastic | 10.38 | 6.11 to 17.64 |
| Neutral | 10.70 | 6.30 to 17.81 |
| Strong clinical benefit |  |  |
| Skeptical | 2.25 | 1.23 to 4.06 |
| Enthusiastic | 2.64 | 1.46 to 4.66 |
| Neutral | 2.80 | 1.57 to 5.00 |
| At least marginal benefit |  |  |
| Skeptical | 5.47 | 3.06 to 9.97 |
| Enthusiastic | 4.39 | 2.41 to 8.17 |
| Neutral | 5.37 | 3.06 to 9.58 |

Abbreviations: aOR, adjusted odds ratio; 95% CrI, 95% credible interval; MCID, minimum clinically important difference.

**Table S3.** Re-analysis of primary outcomes to compute Bayesian posterior probabilities. Four hypotheses were evaluated: the probability of achieving the minimum clinically important difference (MCID) with the experimental arm, defined as hazard ratio (HR) <0.8; the probability of achieving any benefit with the experimental arm, defined as HR <1; the probability of achieving strong clinical benefit with the experimental arm, defined as HR <0.5; and the probability of harm, HR >1. Trials are grouped by the trialist interpretation of either positive or indeterminate/negative, and the median and IQR of the posterior probabilities for included trials are shown.

| **Hypothesis by prior** | **All trials** | **According to trialist interpretation, n (%)** | |
| --- | --- | --- | --- |
|  |  | Positive | Negative |
|  | Median (IQR) | Median (IQR) | Median (IQR) |
| MCID |  |  |  |
| Skeptical | 0.41 (0.05 to 0.94) | 0.92 (0.64 to 1.00) | 0.05 (0.003 to 0.15) |
| Enthusiastic | 0.56 (0.09 to 0.97) | 0.97 (0.76 to 1.00) | 0.08 (0.01 to 0.24) |
| Neutral | 0.49 (0.07 to 0.97) | 0.96 (0.72 to 1.00) | 0.06 (0.005 to 0.18) |
| Strong clinical benefit |  |  |  |
| Skeptical | 0 (0 to 0.09) | 0.01 (0 to 0.22) | 0 (0 to 0) |
| Enthusiastic | 0 (0 to 0.04) | 0.02 (0 to 0.35) | 0 (0 to 0) |
| Neutral | 0 (0 to 0.05) | 0.02 (0 to 0.45) | 0 (0 to 0) |
| At least marginal benefit |  |  |  |
| Skeptical | 0.97 (0.70 to 1.00) | 1.00 (0.99 to 1.00) | 0.67 (0.32 to 0.89) |
| Enthusiastic | 0.99 (0.78 to 1.00) | 1.00 (1.00 to 1.00) | 0.77 (0.49 to 0.94) |
| Neutral | 0.98 (0.72 to 1.00) | 1.00 (1.00 to 1.00) | 0.68 (0.35 to 0.90) |
| Harm |  |  |  |
| Skeptical | 0.03 (0.0002 to 0.30) | 0 (0 to 0.01) | 0.33 (0.11 to 0.66) |
| Enthusiastic | 0.01 (0 to 0.22) | 0 (0 to 0) | 0.23 (0.06 to 0.51) |
| Neutral | 0.02 (0 to 0.28) | 0 (0 to 0) | 0.32 (0.10 to 0.65) |

Abbreviations: MCID, minimum clinically important difference; IQR, interquartile range.

### **Table S4**. List of confounders for each predictor variable identified in the directed acyclic graph depicted in **Figure S5**.

| **Predictor** | **Confounder(s)** |
| --- | --- |
| Disease stage | Disease site |
| Disease site | None identified |
| Cooperative group sponsorship | Disease stage and disease site |
| Industry sponsorship | Disease stage and disease site |
| Enrollment | Cooperative group sponsorship, industry sponsorship, primary endpoint type |
| Year of publication | Enrollment and primary endpoint type |
| Primary endpoint type | Industry sponsorship, cooperative group sponsorship |

**Table S5.** The association of discordance between positive trials and posterior probability of achieving the minimum clinically important difference, HR < 0.8, computed under the enthusiastic prior ≤ 90% by using Bayesian logistic regression, computed with the skeptical prior, and adjusted for confounders selected from the directed acyclic graph in **Figure S5**.

| **Characteristic** | **aOR** | **95% CrI** |
| --- | --- | --- |
| Disease Stage |  |  |
| Solid M0 | 1.05 | 0.59 to 1.88 |
| Solid M1 | Ref |  |
| Hematologic | 0.79 | 0.44 to 1.42 |
| Disease Site |  |  |
| Breast | 0.99 | 0.56 to 1.72 |
| Gastrointestinal | 1.15 | 0.64 to 2.10 |
| Genitourinary | 1.04 | 0.58 to 1.88 |
| Hematologic | 0.73 | 0.42 to 1.26 |
| Thoracic | 0.88 | 0.49 to 1.58 |
| Other | Ref |  |
| Cooperative Group Study | 1.25 | 0.70 to 2.18 |
| Industry Sponsored | 0.68 | 0.36 to 1.30 |
| Number of Enrolled Patients | 1.00 | 1.00 to 1.00 |
| Publication Year | 1.04 | 0.92 to 1.17 |
| Primary Endpoint |  |  |
| Overall Survival | 2.05 | 1.21 to 3.49 |
| Surrogate | Ref |  |

Abbreviations: aOR, adjusted odds ratio; 95% CrI, 95% credible interval; MCID, minimum clinically important difference.

**Table S6**. Subgroup analysis for trials with a primary endpoint of overall survival. The probability of achieving the minimum clinically important difference (MCID) with the experimental arm, defined as hazard ratio (HR) < 0.8; the probability of achieving any benefit with the experimental arm, defined as HR < 1; and the probability of achieving strong clinical benefit with the experimental arm, defined as HR < 0.5, are listed.

| **Hypothesis by prior** | **Posterior probability** | **Trialist interpretation, n (%)** | |
| --- | --- | --- | --- |
|  |  | **Positive (n=30)** | **Negative (n=60)** |
| MCID (HR <0.8) |  |  |  |
| Skeptical | >90% | 8 (27%) | 0 (0%) |
| Skeptical | >75% | 11 (36%) | 0 (0%) |
| Skeptical | >50% | 22 (73%) | 1 (2%) |
| Uninformative | >90% | 9 (30%) | 0 (0%) |
| Uninformative | >75% | 13 (43%) | 0 (0%) |
| Uninformative | >50% | 23 (77%) | 5 (8%) |
| Enthusiastic | >90% | 9 (30%) | 0 (0%) |
| Enthusiastic | >75% | 14 (47%) | 1 (2%) |
| Enthusiastic | >50% | 23 (77%) | 5 (8%) |
| Strong benefit (HR <0.5) |  |  |  |
| Skeptical | >90% | 0 (0%) | 0 (0%) |
| Skeptical | >75% | 0 (0%) | 0 (0%) |
| Skeptical | >50% | 0 (0%) | 0 (0%) |
| Uninformative | >90% | 0 (0%) | 0 (0%) |
| Uninformative | >75% | 0 (0%) | 0 (0%) |
| Uninformative | >50% | 0 (0%) | 0 (0%) |
| Enthusiastic | >90% | 0 (0%) | 0 (0%) |
| Enthusiastic | >75% | 0 (0%) | 0 (0%) |
| Enthusiastic | >50% | 0 (0%) | 0 (0%) |
| At least marginal benefit (HR <1) | |  |  |
| Skeptical | >90% | 30 (100%) | 12 (20%) |
| Skeptical | >75% | 30 (100%) | 24 (40%) |
| Skeptical | >50% | 30 (100%) | 39 (65%) |
| Uninformative | >90% | 30 (100%) | 11 (18%) |
| Uninformative | >75% | 30 (100%) | 24 (40%) |
| Uninformative | >50% | 30 (100%) | 38 (63%) |
| Enthusiastic | >90% | 30 (100%) | 17 (28%) |
| Enthusiastic | >75% | 30 (100%) | 30 (50%) |
| Enthusiastic | >50% | 30 (100%) | 44 (73%) |

Abbreviations: MCID, minimum clinically important difference; HR, hazard ratio.

### **Table S7.** Summary statistics of posterior probabilities according to hypothesis and prior for trials with a primary endpoint of overall survival.

| **Hypothesis by prior** | **Trialist interpretation, n (%)** | |
| --- | --- | --- |
|  | Positive | Negative |
|  | Median (IQR) | Median (IQR) |
| MCID |  |  |
| Skeptical | 0.59 (0.45 to 0.91) | 0.03 (0 to 0.10) |
| Enthusiastic | 0.73 (0.58 to 0.96) | 0.05 (0.01 to 0.19) |
| Neutral | 0.68 (0.53 to 0.94) | 0.04 (0.01 to 0.13) |
| Strong clinical benefit |  |  |
| Skeptical | 0 (0 to 0) | 0 (0 to 0) |
| Enthusiastic | 0 (0 to 0) | 0 (0 to 0) |
| Neutral | 0 (0 to 0) | 0 (0 to 0) |
| At least marginal benefit |  |  |
| Skeptical | 0.99 (0.98 to 1.00 | 0.65 (0.38 to 0.85) |
| Enthusiastic | 1.00 (0.99 to 1.00) | 0.75 (0.46 to 0.92) |
| Neutral | 1.00 (0.99 to 1.00) | 0.65 (0.38 to 0.86) |

Abbreviations: MCID, minimum clinically important difference; IQR, interquartile range.

**Table S8.** Multivariable Bayesian logistic regression between trial-level covariates and discordance for negative trials. Discordance was defined by a negative trial interpretation and probability of HR < 1 exceeding 90% in a Bayesian Cox regression with a skeptical prior. A skeptical prior was used for the logistic regressions.

| **Characteristic** | **aOR** | **95% CrI** |
| --- | --- | --- |
| Disease Stage |  |  |
| Solid M0 | 0.91 | 0.53 to 1.58 |
| Solid M1 | Ref |  |
| Hematologic | 0.95 | 0.49 to 1.86 |
| Disease Site |  |  |
| Breast | 1.16 | 0.64 to 2.10 |
| Gastrointestinal | 1.05 | 0.58 to 1.88 |
| Genitourinary | 1.13 | 0.61 to 2.08 |
| Hematologic | 0.95 | 0.49 to 1.82 |
| Thoracic | 0.80 | 0.44 to 1.45 |
| Other | Ref |  |
| Cooperative Group Study | 1.02 | 0.58 to 1.80 |
| Industry Sponsored | 1.19 | 0.64 to 2.16 |
| Number of Enrolled Patients | 1.00 | 1.00 to 1.00 |
| Publication Year | 0.83 | 0.69 to 0.98 |
| Primary Endpoint |  |  |
| Overall Survival | 1.05 | 0.59 to 1.92 |
| Surrogate | Ref |  |

Abbreviations: M0, non-metastatic; M1, metastatic; aOR, adjusted odds ratio; 95% CrI, 95% credible interval; Ref, reference.

**Table S9**. Correlation between posterior outputs based on individual patient-level data under conventional priors versus summary statistics under the phase III, oncology-specific prior. Concordance was assessed by Lin’s concordance correlation coefficient (ρ_c_).

| **Posterior probability** | **Skeptical prior** | **Enthusiastic prior** | **Neutral prior** |
| --- | --- | --- | --- |
|  | ρ_c_ (95% CI) | ρ_c_ (95% CI) | ρ_c_ (95% CI) |
| At least marginal benefit | 0.97 (0.96 to 0.98) | 0.95 (0.93 to 0.96) | 0.97 (0.96 to 0.98) |
| MCID | 0.98 (0.98 to 0.99) | 0.96 (0.95 to 0.97) | 0.97 (0.97 to 0.98) |
| Strong clinical benefit | 0.98 (0.97 to 0.98) | 0.99 (0.98 to 0.99) | 0.98 (0.97 to 0.98) |
